# Fine Motor Skill Decline after Brain RT – A Multivariate Normal Tissue Complication Probability Study of a Prospective Trial

**DOI:** 10.1101/2022.09.02.22279544

**Authors:** Michael Connor, Mia Salans, Roshan Karunamuni, Soumya Unnikrishnan, Minh-Phuong Huynh-Le, Michelle Tibbs, Alexander Qian, Anny Reyes, Alena Stasenko, Carrie McDonald, Vitali Moiseenko, Issam El-Naqa, Jona Hattangadi-Gluth

## Abstract

**Purpose:** Brain radiotherapy can impair fine motor skills (FMS). FMS are essential for activities of daily living, enabling hand-eye coordination for manipulative movements. We developed normal tissue complication probability (NTCP) models for decline in FMS after fractionated brain RT.

**Methods:** On a prospective trial, 44 primary brain tumor patients received fractioned RT, underwent high-resolution volumetric MRI and diffusion tensor imaging, and comprehensive FMS assessments (Delis-Kaplan Executive Function System Trail Making Test Motor Speed [DKEFS-MS], and Grooved Pegboard Dominant/Non-Dominant Hands [PDH/PNDH]) at baseline and 6 months post-RT. Regions of interest subserving motor function (including cortex, superficial white matter, thalamus, basal ganglia, and white matter tracts) were autosegmented using validated methods and manually verified. Dosimetric and clinical variables were included in multivariate NTCP models, using automated bootstrapped logistic regression, least absolute shrinkage and selection operator (LASSO) logistic regression, and random forests with nested cross-validation.

**Results:** Half of patients showed decline on PNDH, 17 of 42 (40.4%) on PDH, and 11 of 44 (25%) on DKEFS-TM. Automated bootstrapped logistic regression selected a one-term model including maximum dose to dominant postcentral white matter. LASSO selected this term and steroid use. The top five variables in random forest were all dosimetric: mean and maximum dose to dominant corticospinal tract; maximum dose to dominant thalamus; mean dose to dominant caudate; maximum dose to dominant postcentral white matter. This technique performed best with AUC 0.69 (95% CI 0.68 – 0.70) on nested cross-validation.

**Conclusion:** We present the first NTCP models for FMS impairment after brain RT. Dose to several supratentorial motor-associated ROIs correlated with decline in dominant hand fine motor dexterity in primary brain tumor patients, outperforming clinical variables. These data can guide prospective fine motor-sparing strategies for brain RT.

## Introduction

Intracranial radiotherapy (RT) is associated with neurocognitive decline, likely due to injury to eloquent neuroanatomic structures^1^. As survival for brain tumor patients improves, the burden of treatment sequelae increases, with many of these patients exhibiting dysfunction that is progressive and disabling^2^. Although affected neurocognitive domains such as learning, memory, processing speed, attention, and executive function often garner the most interest^3^, fine motor skills (FMS) can also be impaired, with randomized trials demonstrating decline in this domain after RT^4–6^.

FMS involve precise hand-eye coordination for manipulative movements and are essential for many activities of daily living, including handwriting, typing, eating, and using a phone. In addition, FMS are important for performance on other tests of cognitive function^7^. With increased focus on survivorship and quality of life, strategies to improve or preserve these outcomes may consider FMS as well as higher-order cognitive domains. Novel imaging and image processing techniques allow us to well demarcate eloquent brain regions quickly and reliably over time and understand how radiation dose-related damage may underlie functional impairments^4,8–11^.

While neurocognitive decline is a common sequalae of RT for brain tumor patients, there is little in the way of predictive models to guide preservation strategies in radiotherapy patients. We previously reported seminal work on changes in FMS after brain RT, examining in vivo imaging biomarkers of white matter integrity and cortical atrophy in motor-associated regions of interest as predictors of longitudinal FMS decline^4^. This study established associations between radiation dose and loss of cortical and white matter integrity, as well as associations between imaging biomarkers of injury and functional FMS impairment. While we successfully demonstrated the associations between RT dose and microstructural damage within the motor cortex and superficial white matter, normal tissue complication probability (NTCP) analyses would be needed for a more practical, clinical model directly exploring the associations between RT dose and functional outcome. These are the very dose/volume/structure parameters needed to design fine motor functional-sparing interventions. Very few prior reports, mostly pertaining to the hippocampus^12^, have described an NTCP dose response model for a cognitive or functional outcome after brain RT, and QUANTEC (Quantitative Analysis of Normal Tissue Effects in the Clinic) guidelines do not include cognitive or motor outcomes. We sought to perform multivariate NTCP analyses to identify predictors of FMS decline at 6-months post-RT in primary brain tumor patients undergoing fractionated partial brain RT, using data from a prospective clinical trial. Our methodology uses advanced image processing and a cognitive/fine motor neuroscience framework. Given the multiple regions which subserve motor function, and the multiple clinical variables which can affect function, we used multivariate model building techniques. We specifically analyzed baseline to 6-month change in FMS as this approximates the shift from subacute to long-term, irreversible RT-associated damage^2,8^.

## Methods

### Study Overview

We enrolled 52 patients on a single-institution prospective longitudinal IRB-approved clinical trial investigating comprehensive neurocognitive functioning in several domains, including FMS, in patients receiving fractionated RT for primary brain tumors. Patients underwent high resolution volumetric MRI and diffusion imaging, as well as comprehensive neurocognitive evaluation, at baseline (pre-RT) and 3-, 6-, and 12 months after RT completion. All subjects provided written informed consent. Inclusion criteria included: Karnofsky performance status [KPS] ≥70; diagnosis of primary brain tumor; life expectancy of ≥1 year; ability to complete neurocognitive assessments in English; and age ≥18 years. Patients who received prior radiation were excluded. The current analysis includes 44 of these primary brain tumor patients with baseline and 6-month FMS outcomes as well as baseline imaging with the specific MRI protocol detailed below.

### FMS Assessment

FMS was evaluated using two robust, validated objective measurements. All FMS assessments were performed in-person, with direct observation and testing by a trained neuropsychologist. Fine motor speed was evaluated with the Delis-Kaplan Executive Functioning System Trail Making Test Motor Speed^13^ (DKEFS-MS) and fine motor coordination/dexterity was evaluated with the Grooved Pegboard^14^ test. In the DKEFS-MS test, participants are instructed to connect a series of circles joined by a dashed line as quickly as possible. Scores are defined by the time taken to complete the test in seconds. The Grooved Pegboard requires patients to insert metal pegs into slots in sequence as quickly as possible. The score is the time in seconds required to complete the array with the dominant (PDH) and non-dominant (PNDH) hands, with a higher score indicating worse performance.

### Reliable Change Indices

Reliable change indices (RCI), quantifications of whether the change in FMS scores per test is significant for individual patients, were calculated between baseline and 6-month scores using the standardized neurocognitive scores (T-scores)^15^. To account for repeated testing, reliable change indices were adjusted for practice effects (RCI-PEs) using DKEFS-MS T-scores, corrected for age, sex, and education when appropriate, and raw PDH and PNDH scores. RCI-PEs were calculated for each patient on each neurocognitive test measure between 0 and 6 months. RCI-PE calculation is based on the test-retest data of a reference group that has completed the same assessment multiple times^16,17^. A negative RCI-PE was scored as decline.

### Imaging

The imaging acquisition of high-resolution volumetric and diffusion-weighted MRIs for this study have been described in detail elsewhere^18,19^. Briefly, imaging for all patients at each time point were acquired on a 3.0T 750 GE system (GE Healthcare, Milwaukee, Wisconsin) equipped with an 8-channel head coil. Sequences selected for the protocol included a 3D volumetric T1-weighted inversion recovery spoiled gradient echo sequence (echo time [TE]/repetition time [TR]=2.8/6.5 ms; inversion time [TI]=450 ms; flip angle=8 degrees; field of view [FOV]=24 cm), a 3D FLAIR sequence (TE/TR=125/6000 ms, TI=1868 ms, FOV=24cm, matrix=256×256, slice thickness=1 mm), and a diffusion weighted imaging (DWI) sequence using a single-shot pulsed-field gradient spin EPI sequence (TE/TR=96 ms/17 s; FOV=24 cm, matrix=128×128×48; in-plane resolution 1.87×1.875; slice thickness=2.5 mm; 48 slices) with b=0, 500, 1500, and 4000 s/mm^2^, with 1, 6, 6, and 15 unique gradient directions for each b-value respectively and one average for each non-zero b-value. Two additional b=0 volumes were acquired with either forward or reverse phase-encode polarity for use in nonlinear B_0_ distortion correction^20^.

### Image Analysis and Segmentation

Imaging data was preprocessed using in-house algorithms in MATLAB. T1- and T2-weighted images were corrected for distortions attributed to gradient nonlinearities^21^ and imaging non-uniformities arising from bias fields^22^. Diffusion-weighted data was corrected for spatial distortions arising from eddy currents, and B0 field inhomogeneities using robust, well-validated methods^20^.

We specifically investigated brain regions which subserve FMS. As described previously^4^, selected FMS regions of interest (ROIs) included the sensorimotor cortex and superficial white matter (WM) (precentral, postcentral, and paracentral gyri), the corticospinal tracts, the cerebellar cortex and WM, the thalamus, and the basal ganglia (caudate, putamen, and pallidum) (Table S1). ROIs were classified as dominant or non-dominant side per subject based on handedness. For example, in a right-handed patient, the left corticospinal tract was the dominant one. Cortical, superficial WM, cerebellar, thalamic, and basal ganglia ROIs were segmented using the automated FreeSurfer processing pipeline (version 5.3; http://surfer.nmr.harvard.edu), available on the Neuroscience Gateway Portal^23^. Superficial WM is defined as the volume of WM up to 5 mm below the surface of cortical ROIs defined by FreeSurfer’s Desikan-Killiany atlas^24^. The DWI-derived maps and high-resolution volumetric MRI were co-registered and atlas-based tractography was used to segment the DWI into WM tracts in order to evaluate the corticospinal tracts^25^. A censoring mask was drawn manually, slice by slice, on each image to exclude tissue affected by tumor, surgical cavity, or edema. Voxels within the censoring mask were excluded from the final ROI to avoid confounding by tumor and edema-related effects^26^. Planning CT and RT dose maps were co-registered to the baseline T1 and DWI volumes to estimate dose distributions for each of the ROIs^19^.

### Candidate Variables

The mean and maximum doses to all of the above ROIs (Table S1) were calculated and included as potential variables. Volumetric dose variables (eg. V10Gy) were included in univariate analyses, however they were excluded from multivariate model building due to the small volume of most ROIs and high collinearity^27,28^. Additionally, several clinical and demographic variables were included, all binary or continuous: age (years), prescription dose (Gy), receipt of adjuvant chemotherapy (yes/no), anti-epileptic drug use (yes/no), diagnosis (glioma vs. other), ethnicity (Hispanic vs. non-Hispanic), handedness (L vs. R), Karnofsky performance status, KPS (>=90 or <=80), laterality (tumor on dominant or non-dominant side), progression at 6 months (yes/no), race (white vs. other), radiation modality (photon vs. proton), seizures (yes/no), sex (M/F), steroid use (yes/no), any surgery, and GTR (gross total resection) at surgery.

### Statistical Analyses

Statistical analysis was done using R^29^ with packages including mlr3^30^, caret^31^, and glmnet^32^.

### Univariate Analyses

Univariate analyses were performed using logistic regression, Spearman’s rank correlation (Rs), non-parametric tests (Wilcoxon rank test for continuous variables, Fisher’s test for categorical variables), and decision stumps. A decision stump is a one-level decision tree, with a single root node connected immediately to the terminal nodes, and makes a decision based on a single input feature.

### Multivariate Analyses

Multivariate model building was performed using three commonly used methods in this space^33^: automated bootstrapped logistic regression with forward selection, LASSO, and random forests. First, in order to minimize issues with collinearity, among variables highly correlated with each other (Pearson’s correlation coefficient larger than 0.85), those with the lower correlation with the outcome were removed.

The automated logistic regression technique has been described previously, first introduced by El Naqa and colleagues^34^. Briefly, the modeling process is done in two steps. First, an optimal model size (or number of variables included) is estimated by automating forward selection over 1000 bootstrapped samples. The average predictive performance on each out of bag sample for each model order is calculated, and the peak performance of this statistic (e.g., Spearman rank correlation) as a function of the number of variables in the model is used to select the optimal model size. Second, models of this optimal order are fit across the held-out folds of a repeated cross-validation (5-folds, 200 repeats), and the most-frequently selected variable set is chosen for the final model.

LASSO, or least absolute shrinkage and selection operator, is a regression technique with a penalty parameter. Specifically, L1 regularization adds a penalty equal to the absolute value of the coefficients, where lambda is a tuning parameter controlling the amount of regularization. LASSO therefore performs feature selection, encourages sparsity of model parameters, and is well-suited for multicollinearity.

Random Forests are ensembles of many decision trees; the algorithm utilizes both bagging and random subsets of features to create an uncorrelated “forest” of these trees, reducing overfitting. The tunable hyperparameter was the number of variables considered as candidate splitting variables at each split when building each tree, or mtry in the randomForest R package^35^. Variables were ranked by mean decrease in the Gini index; the higher the value the higher the importance of the variable in the model^33^.

Nested cross validation with a 5-fold inner loop for tuning hyperparameters was done; the outer loop also consisting of 5-folds with the held-out data used as a test set for unbiased performance estimation^27^. In each inner loop for the LASSO, an initial feature selection of the top 4 variables by AUC using each feature separately for thresholded class prediction^36^ was also implemented to further reduce the feature space and tendency for overfitting, due to poor performance noted without this. Outer cross-validation with random stratified reshuffling was repeated 200 times. Final models were fit by applying optimal hyperparameters and training on the full dataset.

Model performance, discrimination, and calibration were assessed by the area under the ROC curve (AUC), balanced accuracy, Brier’s score, Nagelke’s R2, calibration slopes and intercepts, and Hosmer-Lemeshow tests.

## Results

### Patient Characteristics and FMS Outcomes

Patient characteristics are shown in Table 1. In total, 44 primary brain tumor patients were eligible for inclusion in this study with baseline and 6-month FMS outcomes and baseline volumetric and diffusion imaging. Of these, all 44 completed baseline and six-month DKEFS-TM tests, and 42 completed pegboard tests. The median age was 46.2 years, 57% were male, and 89% were right-handed. The cohort was high-functioning with 93% having a KPS ≥ 90. Most patients (61%) had gliomas, and 13 (29.5%) had benign diagnoses. By RCI-PE, 21 of 42 patients (50%) experienced decline on the PNDH test at 6 months, 11 of 44 (25%) experienced decline on the DKEFS-TM test, and 17 of 42 (40.4%) declined on the PDH.

**Table 1.**
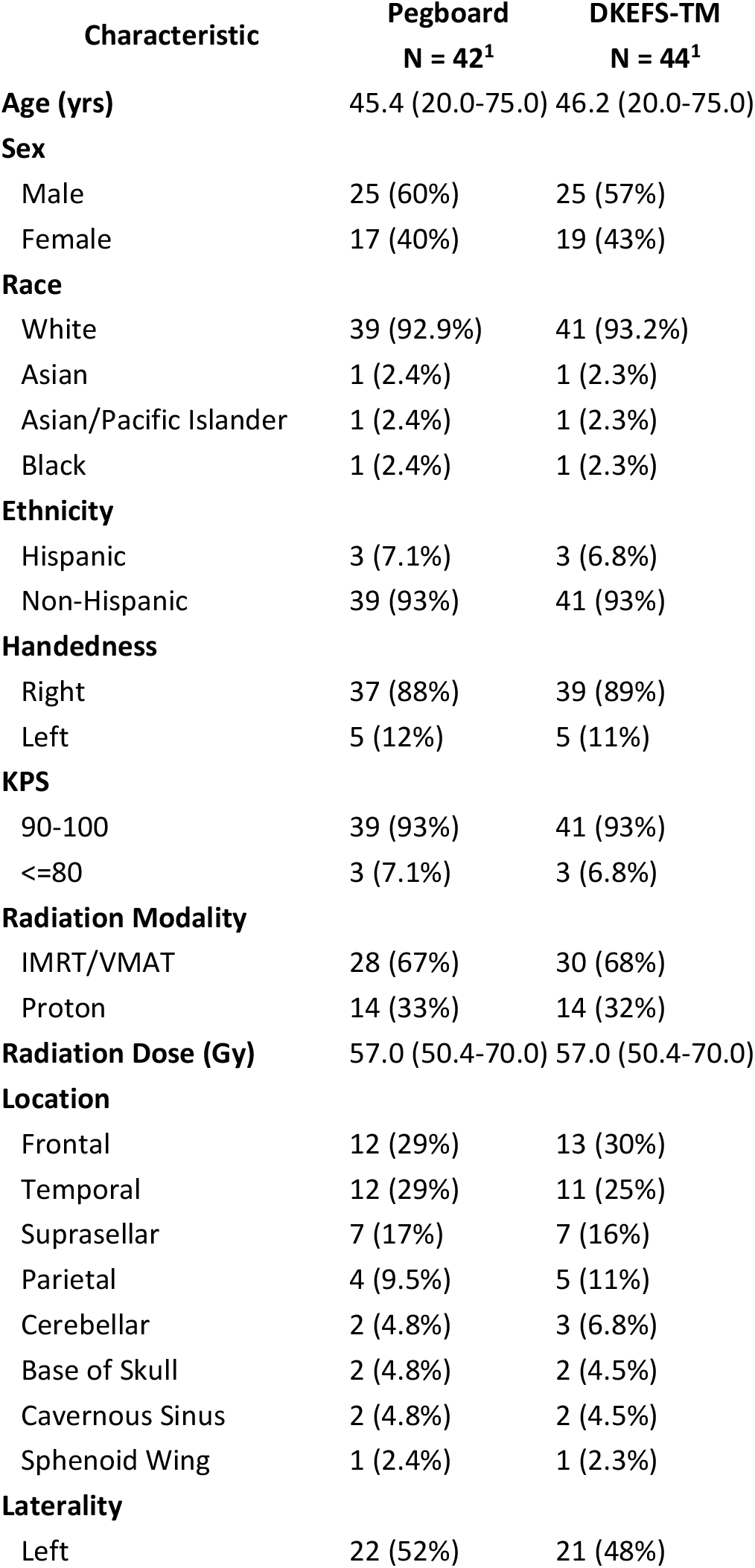
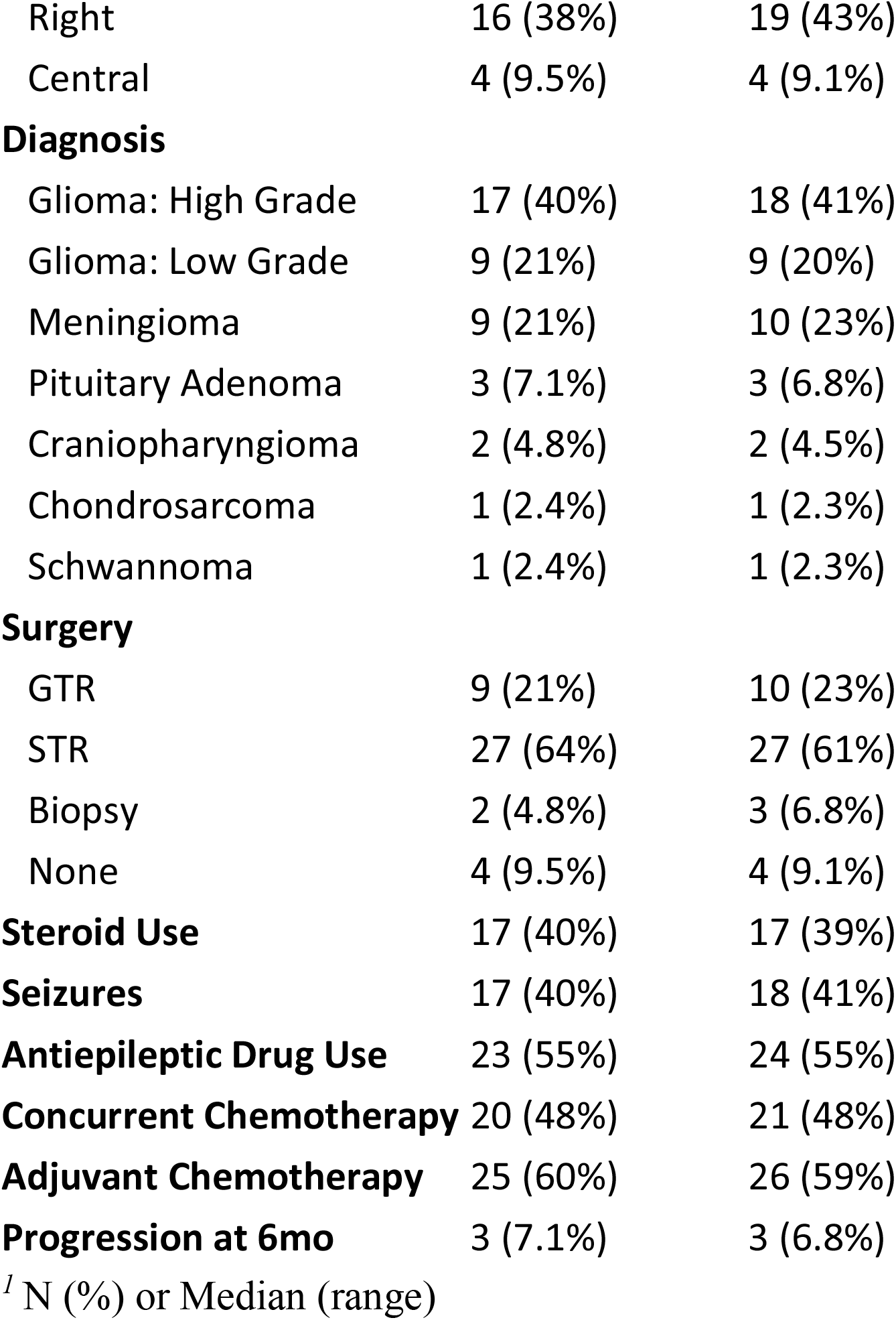
Patient Characteristics.

| **Characteristic** | **Pegboard**<br>**N = 42<sup>1</sup>** | **DKEFS-TM**<br>**N = 44<sup>1</sup>** |
| --- | --- | --- |
| **Age (yrs)** | 45.4 (20.0-75.0) | 46.2 (20.0-75.0) |
| **Sex** |  |  |
| Male | 25 (60%) | 25 (57%) |
| Female | 17 (40%) | 19 (43%) |
| **Race** |  |  |
| White | 39 (92.9%) | 41 (93.2%) |
| Asian | 1 (2.4%) | 1 (2.3%) |
| Asian/Pacific Islander | 1 (2.4%) | 1 (2.3%) |
| Black | 1 (2.4%) | 1 (2.3%) |
| **Ethnicity** |  |  |
| Hispanic | 3 (7.1%) | 3 (6.8%) |
| Non-Hispanic | 39 (93%) | 41 (93%) |
| **Handedness** |  |  |
| Right | 37 (88%) | 39 (89%) |
| Left | 5 (12%) | 5 (11%) |
| **KPS** |  |  |
| 90-100 | 39 (93%) | 41 (93%) |
| <=80 | 3 (7.1%) | 3 (6.8%) |
| **Radiation Modality** |  |  |
| IMRT/VMAT | 28 (67%) | 30 (68%) |
| Proton | 14 (33%) | 14 (32%) |
| **Radiation Dose (Gy)** | 57.0 (50.4-70.0) | 57.0 (50.4-70.0) |
| **Location** |  |  |
| Frontal | 12 (29%) | 13 (30%) |
| Temporal | 12 (29%) | 11 (25%) |
| Suprasellar | 7 (17%) | 7 (16%) |
| Parietal | 4 (9.5%) | 5 (11%) |
| Cerebellar | 2 (4.8%) | 3 (6.8%) |
| Base of Skull | 2 (4.8%) | 2 (4.5%) |
| Cavernous Sinus | 2 (4.8%) | 2 (4.5%) |
| Sphenoid Wing | 1 (2.4%) | 1 (2.3%) |
| **Laterality** |  |  |
| Left | 22 (52%) | 21 (48%) |
| Right | 16 (38%) | 19 (43%) |
| Central | 4 (9.5%) | 4 (9.1%) |
| **Diagnosis** |  |  |
| Glioma: High Grade | 17 (40%) | 18 (41%) |
| Glioma: Low Grade | 9 (21%) | 9 (20%) |
| Meningioma | 9 (21%) | 10 (23%) |
| Pituitary Adenoma | 3 (7.1%) | 3 (6.8%) |
| Craniopharyngioma | 2 (4.8%) | 2 (4.5%) |
| Chondrosarcoma | 1 (2.4%) | 1 (2.3%) |
| Schwannoma | 1 (2.4%) | 1 (2.3%) |
| **Surgery** |  |  |
| GTR | 9 (21%) | 10 (23%) |
| STR | 27 (64%) | 27 (61%) |
| Biopsy | 2 (4.8%) | 3 (6.8%) |
| None | 4 (9.5%) | 4 (9.1%) |
| **Steroid Use** | 17 (40%) | 17 (39%) |
| **Seizures** | 17 (40%) | 18 (41%) |
| **Antiepileptic Drug Use** | 23 (55%) | 24 (55%) |
| **Concurrent Chemotherapy** | 20 (48%) | 21 (48%) |
| **Adjuvant Chemotherapy** | 25 (60%) | 26 (59%) |
| **Progression at 6mo** | 3 (7.1%) | 3 (6.8%) |

<sup>1</sup> N (%) or Median (range)

### Univariate Analyses

On univariate analysis for PDH outcome at 6 months, mean or maximum dose to most dominant supratentorial structures, as well as several of their volumetric dose variables (e.g., V30Gy, V40Gy) were correlated with decline (Table 2, Fig 1). Non-dominant structures showed no association. Using decision stumps and in-sample performance, D_max_ to the precentral cortex, D_mean_ to the caudate, and D_mean_ to the thalamus were the most discriminative dosimetric variables (AUC 0.76-0.77), with cutoff doses of 24.3 Gy, 33.7 Gy, and 27.3 Gy, respectively. Among the clinical variables, laterality (presence of tumor on the dominant side) and steroid use were correlated with decline. There were no significant associations for decline on the non-dominant pegboard test (Table S1), and only increasing age was predictive for decline on the DKEFS-TM (Table S2). Dosimetric variables among dominant motor ROIs showed a high degree of correlation (Fig S1).

**Table 2.** Univariate Analyses for Dosimetric and Demographic/Clinical variables and association with decline on PDH at 6 months.

| Table 2 | Decision Stump |  | Logistic Regression |  | Spearman Correlation |  | Non-parametric |
| --- | --- | --- | --- | --- | --- | --- | --- |
|  | Cutoff (Gy) | AUC (95% CI) | OR (95% CI) | p-value | R<sub>s</sub> | p-value | p-value |
| **Corticospinal Tract** |  |  |  |  |  |  |  |
| D<sub>max</sub> | 53.54 | 0.71 (0.57 - 0.85) | 1.05 (1.01 - 1.10) | 0.03 | 0.36 | 0.02 | 0.01 |
| D<sub>mean</sub> | 19.53 | 0.73 (0.59 - 0.87) | 1.05 (1.01 - 1.11) | 0.03 | 0.39 | 0.01 | 0.01 |
| **Precentral Cortex** |  |  |  |  |  |  |  |
| D<sub>max</sub> | 24.27 | 0.76 (0.62 - 0.89) | 1.04 (1.01 - 1.07) | 0.01 | 0.36 | 0.02 | 0.01 |
| D<sub>mean</sub> | 13.47 | 0.74 (0.60 - 0.88) | 1.05 (1.01 - 1.10) | 0.03 | 0.36 | 0.02 | 0.01 |
| **Precentral WM** |  |  |  |  |  |  |  |
| D<sub>max</sub> | 27.09 | 0.75 (0.61 - 0.89) | 1.04 (1.01 - 1.07) | 0.01 | 0.38 | 0.01 | 0.01 |
| D<sub>mean</sub> | 15.40 | 0.74 (0.60 - 0.88) | 1.05 (1.01 - 1.09) | 0.02 | 0.38 | 0.01 | 0.01 |
| **Postcentral Cortex** |  |  |  |  |  |  |  |
| D<sub>max</sub> | 43.80 | 0.71 (0.57 - 0.85) | 1.04 (1.01 - 1.07) | 0.01 | 0.39 | 0.01 | 0.01 |
| D<sub>mean</sub> | 17.70 | 0.70 (0.56 - 0.84) | 1.05 (1.01 - 1.11) | 0.03 | 0.37 | 0.02 | 0.01 |
| **Postcentral WM** |  |  |  |  |  |  |  |
| D<sub>max</sub> | 44.83 | 0.73 (0.59 - 0.87) | 1.04 (1.01 - 1.08) | 0.01 | 0.42 | 0.01 | <.001 |
| D<sub>mean</sub> | 12.09 | 0.72 (0.58 - 0.86) | 1.05 (1.01 - 1.10) | 0.03 | 0.37 | 0.02 | 0.01 |
| **Paracentral Cortex** |  |  |  |  |  |  |  |
| D<sub>max</sub> | 6.71 | 0.72 (0.57 - 0.86) | 1.02 (1.00 - 1.05) | 0.11 | 0.29 | 0.07 | 0.04 |
| D<sub>mean</sub> | 3.84 | 0.69 (0.54 - 0.83) | 1.02 (0.98 - 1.05) | 0.31 | 0.24 | 0.13 | 0.07 |
| **Paracentral WM** |  |  |  |  |  |  |  |
| D<sub>max</sub> | 59.46 | 0.59 (0.49 - 0.68) | 1.02 (1.00 - 1.05) | 0.08 | 0.28 | 0.08 | 0.04 |
| D<sub>mean</sub> | 3.59 | 0.67 (0.52 - 0.81) | 1.02 (0.99 - 1.06) | 0.20 | 0.24 | 0.13 | 0.07 |
| **Caudate** |  |  |  |  |  |  |  |
| D<sub>max</sub> | 58.88 | 0.69 (0.56 - 0.83) | 1.03 (1.00 - 1.06) | 0.07 | 0.30 | 0.06 | 0.03 |
| D<sub>mean</sub> | 33.70 | 0.76 (0.63 - 0.90) | 1.04 (1.01 - 1.08) | 0.02 | 0.34 | 0.03 | 0.02 |
| **Pallidum** |  |  |  |  |  |  |  |
| D<sub>max</sub> | 38.13 | 0.72 (0.58 - 0.86) | 1.03 (1.00 - 1.07) | 0.04 | 0.29 | 0.06 | 0.03 |
| D<sub>mean</sub> | 20.24 | 0.71 (0.56 - 0.85) | 1.03 (1.00 - 1.07) | 0.03 | 0.29 | 0.07 | 0.04 |
| **Putamen** |  |  |  |  |  |  |  |
| D<sub>max</sub> | 40.94 | 0.72 (0.58 - 0.86) | 1.03 (1.00 - 1.07) | 0.03 | 0.30 | 0.06 | 0.03 |
| D<sub>mean</sub> | 22.40 | 0.73 (0.59 - 0.87) | 1.04 (1.01 - 1.07) | 0.02 | 0.31 | 0.05 | 0.02 |
| **Thalamus** |  |  |  |  |  |  |  |
| D<sub>max</sub> | 53.36 | 0.74 (0.60 - 0.88) | 1.02 (1.00 - 1.06) | 0.11 | 0.24 | 0.13 | 0.07 |
| D<sub>mean</sub> | 27.31 | 0.77 (0.63 - 0.90) | 1.04 (1.01 - 1.07) | 0.03 | 0.27 | 0.09 | 0.04 |
| **Cerebellum Cortex** |  |  |  |  |  |  |  |
| D<sub>max</sub> | 43.12 | 0.64 (0.52 - 0.75) | 0.98 (0.95 - 1.01) | 0.26 | -0.15 | 0.35 | 0.83 |
| D<sub>mean</sub> | 17.54 | 0.62 (0.51 - 0.73) | 0.97 (0.90 - 1.03) | 0.32 | -0.08 | 0.60 | 0.70 |
| **Cerebellum WM** |  |  |  |  |  |  |  |
| D<sub>max</sub> | 41.88 | 0.64 (0.52 - 0.75) | 0.98 (0.95 - 1.01) | 0.29 | -0.16 | 0.33 | 0.84 |
| D<sub>mean</sub> | 23.30 | 0.62 (0.51 - 0.73) | 0.98 (0.92 - 1.03) | 0.39 | -0.10 | 0.55 | 0.73 |
| **Demographic/Clinical** |  |  |  |  |  |  |  |
| Adjuvant Chemotherapy | – | 0.59 (0.44 - 0.74) | 2.22 (0.62 - 8.73) | 0.23 | 0.19 | 0.24 | 0.34 |
| Age | 60.50 | 0.64 (0.51 - 0.77) | 1.02 (0.98 - 1.07) | 0.32 | 0.13 | 0.40 | 0.20 |
| Anti-Epileptic Drug Use | – | 0.63 (0.48 - 0.78) | 3.05 (0.86 - 12.13) | 0.09 | 0.26 | 0.09 | 0.12 |
| Concurrent Chemotherapy | – | 0.54 (0.39 - 0.70) | 1.43 (0.42 - 5.04) | 0.57 | 0.09 | 0.58 | 0.75 |
| Diagnosis (Other) | – | 0.57 (0.42 - 0.72) | 0.53 (0.13 - 1.91) | 0.34 | -0.15 | 0.35 | 0.5 |
| Dose | 60.60 | 0.56 (0.48 - 0.64) | 1.14 (0.98 - 1.38) | 0.12 | 0.21 | 0.19 | 0.10 |
| Ethnicity (Non-Hispanic) | – | 0.56 (0.49 - 0.63) | – | – | 0.23 | 0.15 | 0.26 |
| Handedness (R) | – | 0.50 (0.40 - 0.60) | 1.02 (0.15 - 8.48) | 0.98 | <.001 | 0.98 | 1.00 |
| KPS ($\leq 80$) | – | 0.54 (0.45 - 0.63) | 3.20 (0.28 - 72.45) | 0.36 | 0.15 | 0.35 | 0.56 |
| Laterality (Dominant) | – | 0.68 (0.54 - 0.83) | 4.87 (1.31 - 21.49) | 0.02 | 0.36 | 0.02 | 0.03 |
| Progression at 6mo | – | 0.51 (0.43 - 0.59) | 0.72 (0.03 - 8.13) | 0.79 | -0.04 | 0.80 | 1.00 |
| Race (Other) | – | 0.53 (0.44 - 0.62) | 0.46 (0.02 - 3.96) | 0.52 | -0.10 | 0.52 | 0.64 |
| Radiation Modality (Protons) | – | 0.52 (0.37 - 0.67) | 1.16 (0.31 - 4.28) | 0.82 | 0.03 | 0.83 | 1.00 |
| Seizures | – | 0.65 (0.50 - 0.80) | 3.67 (1.03 - 14.24) | 0.05 | 0.31 | 0.05 | 0.06 |
| Sex (M) | – | 0.51 (0.35 - 0.66) | 0.95 (0.27 - 3.41) | 0.94 | -0.01 | 0.94 | 1.00 |
| Steroid Use | – | 0.70 (0.56 - 0.85) | 5.81 (1.57 - 24.21) | 0.01 | 0.41 | 0.01 | 0.01 |
| Surgery (any) | – | 0.52 (0.41 - 0.63) | 0.70 (0.09 - 4.08) | 0.70 | -0.06 | 0.71 | 1.00 |
| Surgery (GTR) | – | 0.52 (0.39 - 0.65) | 0.81 (0.18 - 3.82) | 0.79 | -0.04 | 0.79 | 1.00 |

**Fig 1.**
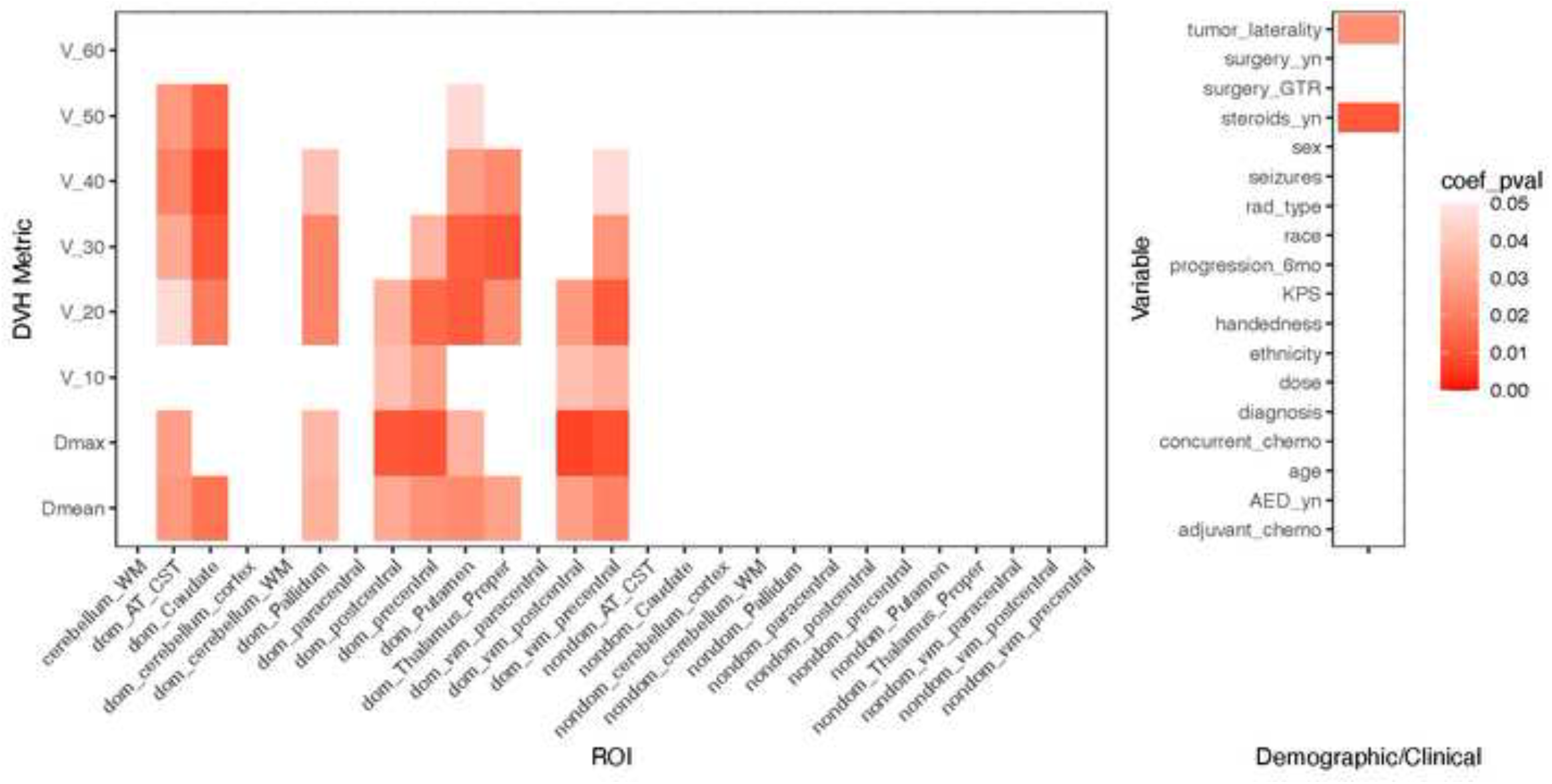
Heatmap for univariate logistic regression coefficient p-values for decline on PDH

### Multivariate Analyses

We proceeded with multivariate model building for decline on the PDH at 6 months. Automated bootstrapped logistic regression selected an optimal model order of 1, likely reflecting overfitting with higher order models due to our small sample size. The most frequently selected variable was maximum dose to the dominant postcentral white matter, with the next two most frequent being steroid use, and mean dose to the dominant caudate (Fig 2A). Model performance at nested cross-validation by the AUC as 0.61 (SD 0.005) (Fig 3). The final NTCP model is given by:

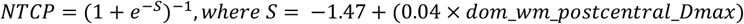

**Fig 2.**
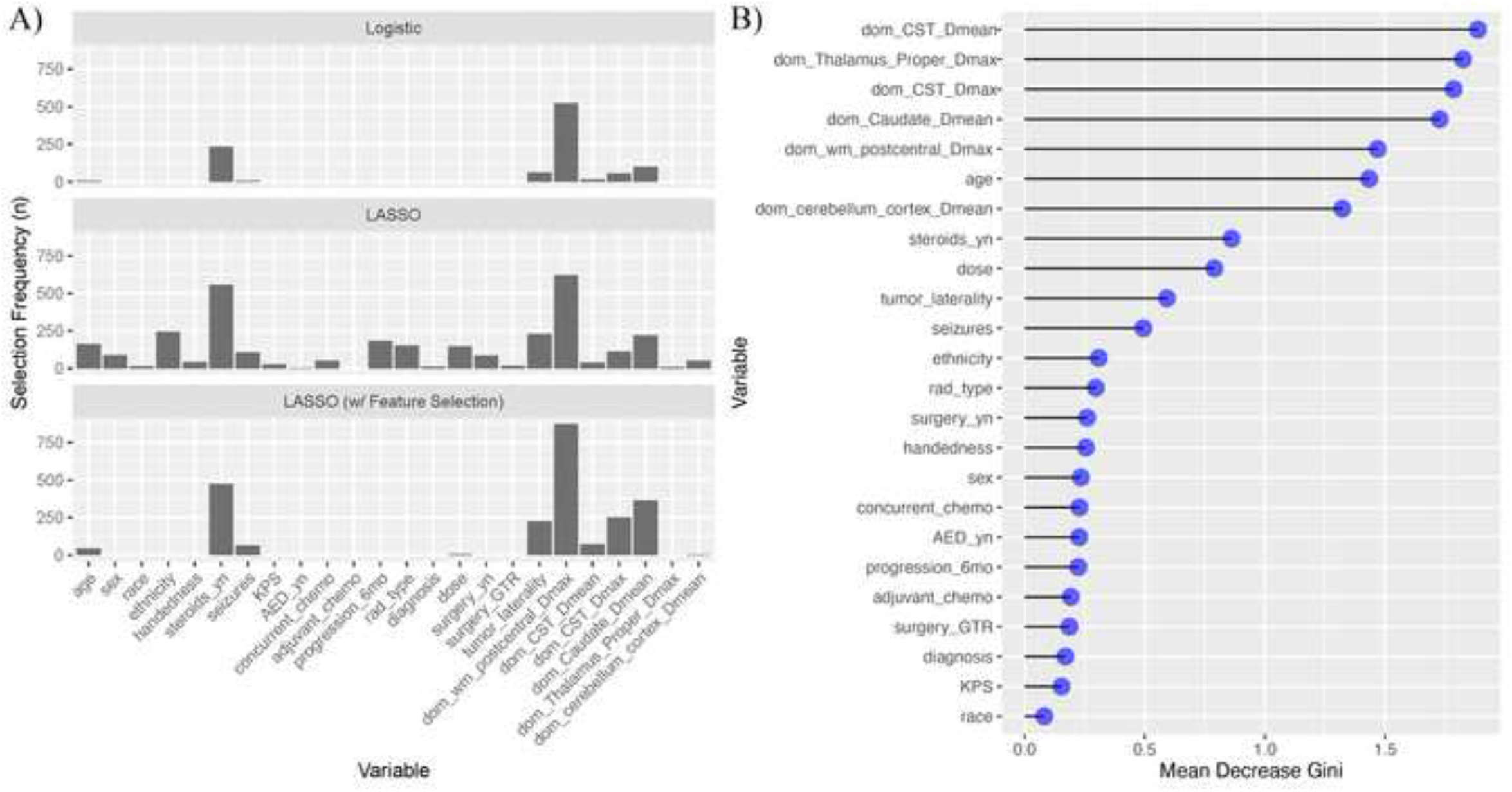
A) Frequency of variable selection over the cross-validation folds for the logistic methods. B) Variable importance in the random forest model by mean decrease in Gini score.

**Fig 3.**
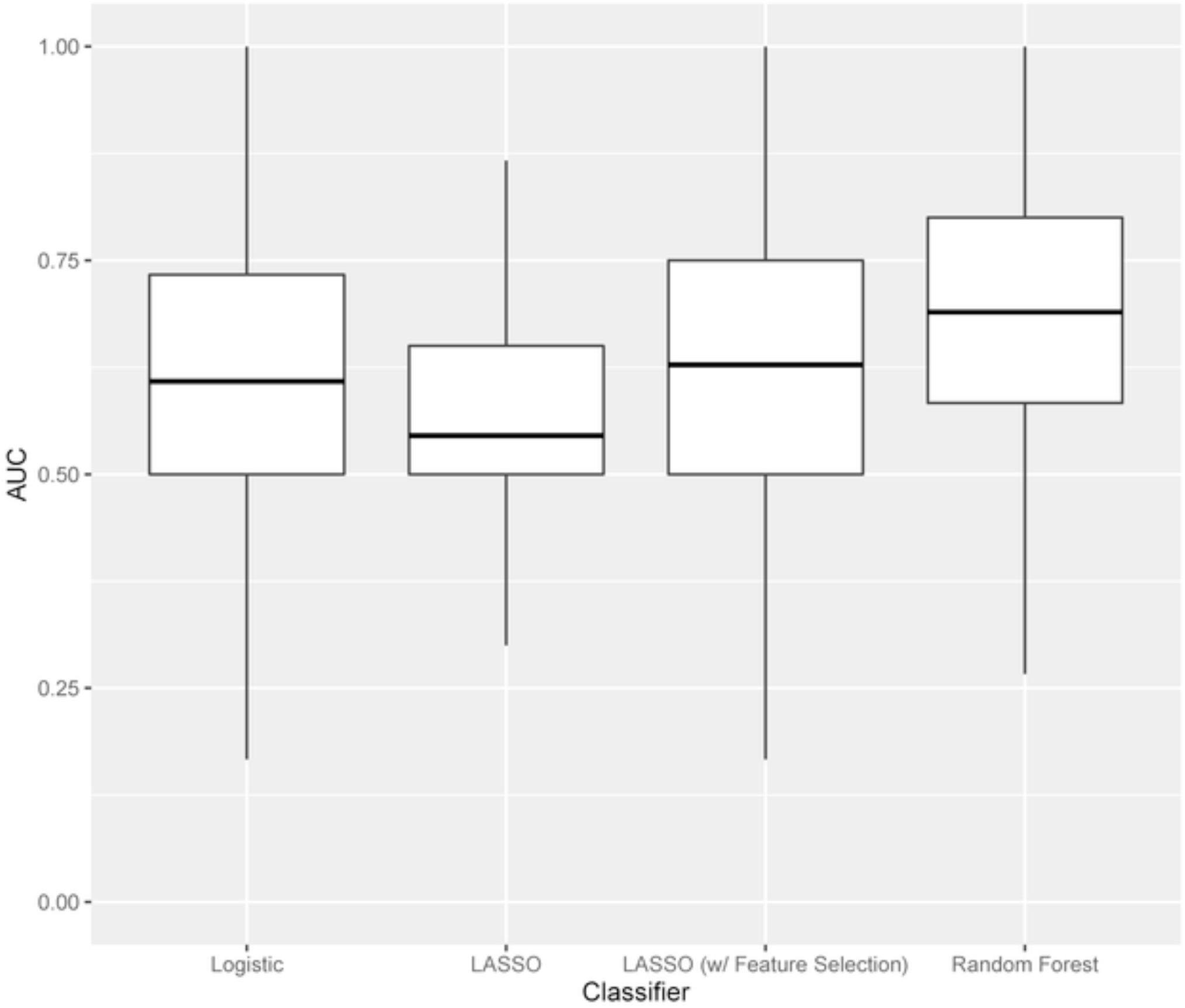
Boxplots of area under the curve (AUC) performance over the repeated outer cross-validation held-out folds.

LASSO model performance at nested cross validation by AUC was 0.55 (SD 0.005), improved to 0.63 (SD 0.005) by the use of an initial feature selection step based on AUC (Fig 3). The frequency of terms selected over the repeated, nested CVs are shown in Fig 2A. The final LASSO NTCP model (without feature selection) is given by:

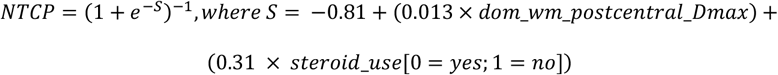

The final LASSO NTCP model (with feature selection) is given by:

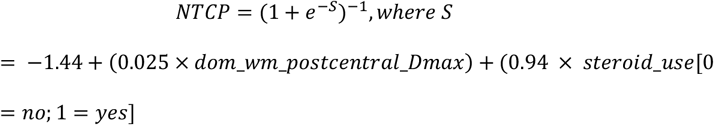

Graphical depictions of the logistic NTCP curves are shown in the supplement (Fig S3, S4).

The top five important variables in the Random Forest, as ranked by mean decrease in Gini coefficient, were: mean dose to dominant corticospinal tract; maximum dose to the dominant thalamus; mean dose to dominant corticospinal tract; mean dose to the dominant caudate, and maximum dose to the dominant postcentral white matter (Fig 2B). All of these dosimetric variables outperformed clinical variables. Model performance by AUC was 0.69 (95% CI 0.68 – 0.70) on nested cross-validation (Fig 3).

All models achieved good calibration by Hosmer-Lemeshow test (p=0.07-0.67). Additional model characteristics are shown in Table 3.

**Table 3.** Performance and characteristics for each model-building method.

|  | Logistic | LASSO | LASSO (w/ FS) | Random Forest |
| --- | --- | --- | --- | --- |
| *<u>Final Fitted Model</u>* |  |  |  |  |
| AUC | 0.75 | 0.79 | 0.80 | 0.70 |
| Balanced Accuracy | 0.71 | 0.71 | 0.76 | 0.68 |
| Brier Score | 0.19 | 0.20 | 0.18 | 0.21 |
| Calibration Slope | 1.00 | 2.89 | 1.39 | 0.79 |
| Calibration Intercept | 0.00 | 0.59 | 0.11 | -0.10 |
| Hosmer–Lemeshow p-value | 0.67 | 0.07 | 0.23 | 0.57 |
| R<sup>2</sup> | 0.25 | 0.22 | 0.34 |  |
| *<u>Nested Cross Validation</u>* |  |  |  |  |
| AUC | 0.61 (SD 0.005) | 0.55 (SD 0.005) | 0.63 (SD 0.005) | 0.69 (SD 0.006) |
| Balanced Accuracy | 0.62 (SD 0.005) | 0.57 (SD 0.003) | 0.62 (SD 0.004) | 0.67 (SD 0.005) |
| Brier Score | 0.26 (SD 0.002) | 0.26 (SD 0.002) | 0.24 (SD 0.002) | 0.23 (SD 0.002) |

Multivariate analysis was attempted for the other endpoints, though models performed poorly via nested cross validation for predicting decline on the PNDH (AUC 0.31-0.49) and DKEFS-TM (AUC 0.51-0.57) tests.

## Discussion

There is a critical need to understand substructure-dose-function relationships in the brain, and to build practical and clinical models for cognitive and functional outcomes, thereby paving the way for more individualized precision brain RT. We present the <u>first evidence-based NTCP modeling analyses of fine motor function decline</u> after brain RT, and the only study, to our knowledge, of a neurocognitive/functional endpoint outside of hippocampal dose and cognition. There are published NTCP studies for the prediction of memory decline based on hippocampal dose^37–39^, however one of these failed an external validation attempt^40^ and none use multivariate methods or take into account clinical variables. Given that the probability of a functional change in outcome is typically influenced by both dosimetric and clinical factors, we employed multivariate model building methods^34^. We also employed a neuroanatomic and motor function framework and advanced image processing to analyze specific brain regions of interest which are associated with fine motor function. We found that dose to several supratentorial motor-associated ROIs correlated with decline on grooved pegboard dominant hand tests at 6 months post-RT in primary brain tumor patients undergoing partial fractionated brain RT. These findings implicate the anatomic-functional pathway of fine motor control: including cortex, superficial white matter, thalamus and basal ganglia, and the corticospinal tract. The results also reinforce neuroanatomic correlation; that dose to brain ROIs on the motor dominant side of the cerebral hemisphere matter most.

Although there is more focus on higher-order cognitive domains in the literature, impairment in fine motor skills has been consistently reported in clinical trials^4–6^. Chang et al found that 33-50% of patients undergoing SRS vs. SRS plus whole brain RT experienced significant decline on grooved pegboard tests of fine motor skills, the highest rates of decline outside of the Hopkins Verbal Learning Test (HVLT) in the study^6^. Brown et al, in reporting the results of Alliance N0574, found similar rates of decline on grooved pegboard tests in patients with 1-3 metastases randomized to SRS with or without whole brain RT, with 29.3-47.7% of patients declining on grooved pegboard tests^5^. Moreover, fine motor skills are critical for performance on other common neurocognitive instruments used in clinical trials, including tests of processing speed and those that require a writing/drawing component. Some investigators have even used fine motor skill tests as a “measure of higher-level cognitive processes which influence motor and nonmotor skills alike”^41–43^.

We previously found that increasing age and use of anti-epileptic medications predicted higher rates of fine motor skill decline among primary brain tumor patients undergoing fractionated RT^4^. We also found imaging biomarkers for FMS decline, showing that diffusion imaging measures of white matter microstructure injury in the cerebellar white matter and corticospinal tract were predictive of decline, as was volumetric atrophy of the precentral and cerebellar cortices, pallidum, and pons^4^. Likewise, in traumatic brain injury, atrophy of the thalamus, putamen, and pallidum have been correlated significantly with decline in FMS^44^. Here, we report the results of NTCP modeling to directly predict fine motor skill decline from planned/delivered doses and clinical variables.

Univariate methods showed that mean or max dose to all supratentorial motor ROIs in our study were associated with FMS decline, with a high degree of correlation among variables. Multivariate approaches yielded very parsimonious models likely due to our modest sample size. Multivariate logistic regression techniques (automated bootstrapped logistic regression and LASSO) selected maximum dose to the dominant postcentral white matter and steroid use as the top two features for inclusion in the models. The top five most important predictors in the Random Forest (Fig 2B) were all dosimetric variables relating to dominant-sided supratentorial motor ROIs, all of which outperformed clinical variables. These results reinforce the importance of dose to motor-associated functional brain areas, with careful attention to lateralization of brain function affecting the dominant hand.

One may expect that dose to the precentral, or primary motor, cortex would be most important in a model predicting fine motor skill decline, and indeed these variables were ranked as highly important in the random forest. Dose variables for the precentral cortex were also significant on univariate analysis (Table 2). Additionally, D_max_ to the precentral cortex was highly predictive as a decision stump. However, our logistic models selected maximum dose to the postcentral white matter and random forest showed high importance of this region. As an illustrative example, the logistic model predicts increased risk of decline on PDH of 5%, 10%, and 20% over baseline with doses of 6.9 Gy, 13.1 Gy, and 25 Gy, respectively, to the postcentral white matter (Fig. S3) This could represent a nuance of our dataset, as dose to this postcentral area is highly collinear with doses to the precentral cortex given they are neuroanatomically adjacent. However, some studies have demonstrated the importance of injury to the somatosensory cortex to motor function and recovery^45,46^. Mean dose to caudate on the dominant side, and max dose to thalamus were consistently important dosimetric predictors in our study. These findings are consistent with previous studies of the basal ganglia and thalamus as critical structures in radiosurgery applications. These two areas ranked behind only the pons/midbrain as high-risk regions in a clinical study to predict permanent symptomatic post-SRS injury in AVM patients^47^. Finally, mean and maximum dose to the dominant corticospinal tract was an important predictor of FMS decline in the dominant hand in our study. This key white matter tract represents a possible (organ at risk) OAR for sparing.

Indeed, there is limited precedent in the literature for implemented motor-associated OARs into treatment planning. Maruyama et al found that a maximum dose of 23 Gy to the corticospinal tract resulted in a 5% complication rate^48^. Koga et al found that limiting the corticospinal tract to 20 Gy during single fraction radiosurgery for AVMs reduced motor complications without decreasing the obliteration rate^49^. We are currently testing motor-associated OAR avoidance, along with avoidance of other eloquent white matter tracts, during SRS on a prospective clinical trial in patients with brain metastases^50^. However, in many instances involving brain tumors, a certain amount of dose to nearby structures is unavoidable. NTCP models may therefore also be used in the future to identify patients at high risk of toxicity, in whom preventative or rehabilitative strategies may be undertaken. The rehabilitation of motor skills has been found to result in persistent gray matter changes over the short term, suggesting this may counteract the atrophy noted in RT and other studies^51^. The restoration of motor skill after injury may involve enhanced activation of contralateral cortex as well^52^.

In terms of clinical variables, patient age emerged as the most important demographic/clinical variable. Other studies have confirmed the finding that increasing age may compromise performance on the grooved pegboard test^4,53,54^, as fine motor dexterity declines with normal and pathological aging. Steroid use emerged as a predictive factor in several of our models for FMS decline. This may be a surrogate marker for tumor volume, edema, mass effect, and/or overall functioning. Prolonged steroid use may also cause myopathy in some patients, though this is usually associated with large muscle groups and not fine motor control. Use of anti-epileptic mediations has also been associated with decline in FMS^55^, however, this was not a particularly important variable in our analyses.

Our study does have potential limitations. Performance on nested cross-validation was considerably lower than the final models fit to all data, suggesting an unavoidable degree of optimism or overfitting given our modest sample size. Nevertheless, our best models attained acceptable discrimination ability (Random Forest, AUC 0.69) on validation, and the models were well-calibrated. The inclusion of several ROIs and several dosimetric quantities with high collinearity necessitates variable selection for parsimonious, interpretable models with optimal bias-variance trade-off. Motor ROIs were segmented using robust methods for parcellation of white matter tracts and cortical and subcortical structures; these methods are well-validated^4,8–11^ in the neuroimaging literature, including in brain tumor patients. To minimize any confounding by tumor, segmentations for each patient were inspected slice by slice, manually censoring tumor, surgical cavities, and edema. The endpoint of decline on fine motor skill tests may be influenced by other biomedical or psychological factors beyond the variables we considered^41^. Our sample size was modest, though we employed a nested cross-validation approach in order to avoid overfitting as much as possible. The current study sample size is similar or larger than other studies on hippocampal dose-response models^12,39,56^. We enrolled a heterogeneous group of brain tumor patients, and tumor type or receipt of chemotherapy were not important variables in our models, though our sample size likely limits these findings. With our heterogeneous group and validation however, our findings are more generalizable to all primary brain tumor patients undergoing intracranial RT. Our models do not exhibit excellent discriminatory power, but perform on par with other recently developed multivariate NTCP models predicting xerostomia^57^, esophagitis^58^, rectal and bladder morbidity^59^, or radiation pneumonitis^60^, for example. Outcomes were prospectively gathered, and objectively measured by a neuropsychologist. Our models rely on a pure structure-function paradigm, which the complexities of the human brain may certainly defy. Network connectivity is a growing area of research utilizing graph theory, borrowed from computer science, where the brain is represented as network of nodes (anatomical areas under consideration) and edges (the interconnections between these areas). Diminished connectivity in motor networks among patients with brain tumors and weakness has been documented^61^, and future studies may consider these more complex networks models to predict motor performance.

In conclusion, we present the first NTCP models for a neurologic functional endpoint outside of hippocampal dose and cognition. Specifically, we found that the dose to several supratentorial motor-associated ROIs correlated with decline on pegboard dominant hand tests at 6 months post-RT in primary brain tumor patients undergoing partial fractionated brain RT. Future studies may externally validate these models and employ prospective strategies to minimize loss of fine motor function after RT.

## Data Availability

All data produced in the present study are available upon reasonable request to the authors

## Acknowledgements

Special thanks to the patients who enrolled and participated in this study. Many thanks to nurses in radiation oncology, particularly Abby Pennington, RN and Mary Kay Gorman, RN, for their help with this study.

## References

1. Greene-Schloesser D, Robbins ME, Peiffer AM, Shaw EG, Wheeler KT, Chan MD. Radiation-induced brain injury: A review. Front Oncol. 2012;2(July):1–18.

2. Makale MT, McDonald CR, Hattangadi-Gluth JA, Kesari S. Mechanisms of radiotherapy-associated cognitive disability in patients with brain tumours. Nat Rev Neurol. 2017;13(1):52–64.

3. Meyers CA, Brown PD. Role and relevance of neurocognitive assessment in clinical trials of patients with CNS tumors. J Clin Oncol. 2006;24(8):1305–1309.

4. XXXX

5. Brown PD, Jaeckle K, Ballman K V, et al. Effect of Radiosurgery Alone vs Radiosurgery With Whole Brain Radiation Therapy on Cognitive Function in Patients With 1 to 3 Brain Metastases: A Randomized Clinical Trial. JAMA. 2016;316(4):401–409.

6. Chang EL, Wefel JS, Hess KR, et al. Neurocognition in patients with brain metastases treated with radiosurgery or radiosurgery plus whole-brain irradiation: a randomised controlled trial. Lancet Oncol. 2009;10(11):1037–1044.

7. Bezdicek O, Nikolai T, Hoskovcová M, et al. Grooved Pegboard Predicates More of Cognitive Than Motor Involvement in Parkinson’s Disease. Assessment. 2014;21(6):723–730.

8. Huynh-Le MP, Tibbs MD, Karunamuni R, et al. Microstructural Injury to Corpus Callosum and Intrahemispheric White Matter Tracts Correlate With Attention and Processing Speed Decline After Brain Radiation. Int J Radiat Oncol Biol Phys. 2021;110(2):337–347.

9. Tibbs MD, Huynh-Le MP, Karunamuni R, et al. Microstructural Injury to Left-Sided Perisylvian White Matter Predicts Language Decline After Brain Radiation Therapy. Int J Radiat Oncol Biol Phys. 2020.

10. Tringale KR, Nguyen TT, Karunamuni R, et al. Quantitative Imaging Biomarkers of Damage to Critical Memory Regions Are Associated With Post–Radiation Therapy Memory Performance in Brain Tumor Patients. Int J Radiat Oncol Biol Phys. 2019;105(4):773–783.

11. Tringale KR, Nguyen T, Bahrami N, et al. Identifying early diffusion imaging biomarkers of regional white matter injury as indicators of executive function decline following brain radiotherapy: A prospective clinical trial in primary brain tumor patients. Radiother Oncol. 2019;132:27–33.

12. Gondi V, Hermann BP, Mehta MP, Tome WA. Hippocampal dosimetry predicts neurocognitive function impairment after fractionated stereotactic radiotherapy for benign or low-grade adult brain tumors. Int J Radiat Oncol Biol Phys. 2012;83(4):e487–93.

13. Delis D, Kaplan E, Kramer J. Delis-Kaplan Executive Function System (D-KEFS). 2001.

14. Lafayette Instrument. Grooved Pegboard Test. 2015.

15. Jacobson NS, Truax P. Clinical Significance: A Statistical Approach to Defining Meaningful Change in Psychotherapy Research. J Consult Chmcal Psychol. 1945;59(1):12–19.

16. Strauss E, Sherman EMS, Spreen O. A Compendium of Neuropsychological Tests: Administration, Norms, and Commentary. 3rd ed. Oxford University Press

17. Chelune GJ, Naugle RI, Lüders H, Sedlak J, Awad IA. Individual change after epilepsy surgery: Practice effects and base-rate information. Neuropsychology. 1993;7(1):41–52.

18. XXXX

19. XXXX

20. Holland D, Kuperman JM, Dale AM. Efficient correction of inhomogeneous static magnetic field-induced distortion in Echo Planar Imaging. Neuroimage. 2010;50(1):175–183.

21. Jovicich J, Czanner S, Greve D, et al. Reliability in multi-site structural MRI studies: Effects of gradient non-linearity correction on phantom and human data. Neuroimage. 2006;30(2):436–443.

22. Sled JG, Zijdenbos AP, Evans AC. A nonparametric method for automatic correction of intensity nonuniformity in MRI data. IEEE Trans Med Imaging. 1998;17(1):87–97.

23. Carnevale T, Majumdar A, Sivagnanam S, et al. The neuroscience gateway portal: high performance computing made easy. BMC Neurosci. 2014;15(S1):1–1.

24. Sivagnanam S, Majumdar A, Yoshimoto K, et al. Introducing the neuroscience gateway. CEUR Workshop Proc. 2013;993.

25. Hagler Jr. DJ, Ahmadi ME, Kuperman J, et al. Automated White-Matter Tractography Using a Probabilistic Diffusion Tensor Atlas : Application to Temporal Lobe Epilepsy. 2009;1547(February 2008):1535–1547.

26. Connor M, Karunamuni R, McDonald C, et al. Regional susceptibility to dose-dependent white matter damage after brain radiotherapy. Radiother Oncol. 2017;123(2):209–217.

27. Xu CJ, Van Der Schaaf A, Van’T Veld AA, Langendijk JA, Schilstra C. Statistical validation of normal tissue complication probability models. Int J Radiat Oncol Biol Phys. 2012;84(1):e123–e129.

28. Cella L, Liuzzi R, Conson M, D’Avino V, Salvatore M, Pacelli R. Multivariate normal tissue complication probability modeling of heart valve dysfunction in hodgkin lymphoma survivors. Int J Radiat Oncol Biol Phys. 2013;87(2):304–310.

29. R Core Team (2015). R: A Language and Environment for Statistical Computing, Vienna, Australia. https://www.r-project.org.

30. Lang M, Binder M, Richter J, et al. mlr3: A modern object-oriented machine learning framework in R. J Open Source Softw. 2019;4(44):1903.

31. Kuhn M. Building predictive models in R using the caret package. J Stat Softw. 2008;28(5):1–26.

32. Friedman J, Hastie T, Tibshirani R. Regularization Paths for Generalized Linear Models via Coordinate Descent. J Stat Softw. 2010;33(1):1–22.

33. Naqa I El. A Guide To Outcome Modeling In Radiotherapy and Oncology. Boca Raton, FL: CRC Press, Taylor and Francis Group; 2018.

34. El Naqa I, Bradley J, Blanco AI, et al. Multivariable modeling of radiotherapy outcomes, including dose–volume and clinical factors. Int J Radiat Oncol. 2006;64(4):1275–1286.

35. Liaw A, Wiener M. Classification and Regression by randomForest. R News. 2002;2(3):18–22.

36. Bommert A, Sun X, Bischl B, Rahnenführer J, Lang M. Benchmark for filter methods for feature selection in high-dimensional classification data. Comput Stat Data Anal. 2020;143:106839.

37. Gondi V, Hermann BP, Mehta MP, Tome WA. Hippocampal dosimetry predicts neurocognitive function impairment after fractionated stereotactic radiotherapy for benign or low-grade adult brain tumors. Int J Radiat Oncol Biol Phys. 2013;85(2):348–354.

38. Ma TM, Grimm J, McIntyre R, et al. A prospective evaluation of hippocampal radiation dose volume effects and memory deficits following cranial irradiation. Radiother Oncol. 2017;125(2):234–240.

39. Acharya S, Wu S, Ashford JM, et al. Association between hippocampal dose and memory in survivors of childhood or adolescent low-grade glioma: A 10-year neurocognitive longitudinal study. Neuro Oncol. 2019;21(9):1175–1183.

40. Jaspers J, Mèndez Romero A, Hoogeman MS, et al. Evaluation of the Hippocampal Normal Tissue Complication Model in a Prospective Cohort of Low Grade Glioma Patients—An Analysis Within the EORTC 22033 Clinical Trial. Front Oncol. 2019;9(October):1–9.

41. Goldstein G, Nussbaum PD, Beers SR. Neuropsychology. 1st ed. New York: Springer Science+Business Media, LLC; 1997.

42. Diamond A. Close interrelation of motor development and cognitive development and of the cerebellum and prefrontal cortex. Child Dev. 2000;71(1):44–56.

43. Tolle KA, Rahman-Filipiak AM, Hale AC, Kitchen Andren KA, Spencer RJ. Grooved Pegboard Test as a measure of executive functioning. Appl Neuropsychol. 2020;27(5):414–420.

44. Gooijers J, Chalavi S, Beeckmans K, et al. Subcortical Volume Loss in the Thalamus, Putamen, and Pallidum, Induced by Traumatic Brain Injury, Is Associated with Motor Performance Deficits. Neurorehabil Neural Repair. 2016;30(7):603–614.

45. Abela E, Missimer J, Wiest R, et al. Lesions to primary sensory and posterior parietal cortices impair recovery from hand paresis after stroke. PLoS One. 2012;7(2).

46. Kato H, Izumiyama M. Impaired motor control due to proprioceptive sensory loss in a patient with cerebral infarction localized to the postcentral gyrus. J Rehabil Med. 2015;47(2):187–190.

47. Flickinger JC, Kondziolka D, Dade Lunsford † L, et al. Development of a Model To Predict Permanent Symptomatic Postradiosurgery Injury For Arteriovenous Malformation Patients. Int J Radiat Oncol Biol Phys. 2000;46(5):1143–1148.

48. Maruyama K, Kamada K, Ota T, et al. Tolerance of Pyramidal Tract to Gamma Knife Radiosurgery Based on Diffusion-Tensor Tractography. Int J Radiat Oncol Biol Phys. 2008;70(5):1330–1335.

49. Koga T, Shin M, Maruyama K, et al. Integration of corticospinal tractography reduces motor complications after radiosurgery. Int J Radiat Oncol Biol Phys. 2012;83(1):129–133.

50. XXXX

51. Filippi M, Ceccarelli A, Pagani E, et al. Motor learning in healthy humans is associated to gray matter changes: A tensor-based morphometry study. PLoS One. 2010;5(4).

52. Schaechter JD, Perdue KL. Enhanced cortical activation in the contralesional hemisphere of chronic stroke patients in response to motor skill challenge. Cereb Cortex. 2008;18(3):638–647.

53. Yao ZF, Yang MH, Hsieh S. Brain Structural-Behavioral Correlates Underlying Grooved Pegboard Test Performance Across Lifespan. J Mot Behav. 2020;0(0):1–12.

54. Nyquist PA, Yanek LR, Bilgel M, et al. Effect of white matter lesions on manual dexterity in healthy middle-aged persons. Neurology. 2015;84(19):1920–1926.

55. Mula M, Trimble MR. Antiepileptic Drug-Induced Cognitive Adverse Effects. CNS Drugs. 2009;23(2):121–137.

56. Okoukoni C, McTyre ER, Ayala Peacock DN, et al. Hippocampal dose volume histogram predicts Hopkins Verbal Learning Test scores after brain irradiation. Adv Radiat Oncol. 2017;2(4):624–629.

57. Teng F, Fan W, Luo Y, et al. A Risk Prediction Model by LASSO for Radiation-Induced Xerostomia in Patients With Nasopharyngeal Carcinoma Treated With Comprehensive Salivary Gland–Sparing Helical Tomotherapy Technique. Front Oncol. 2021;11(February):1–9.

58. Luna JM, Chao HH, Shinohara RT, et al. Machine learning highlights the deficiency of conventional dosimetric constraints for prevention of high-grade radiation esophagitis in non-small cell lung cancer treated with chemoradiation. Clin Transl Radiat Oncol. 2020;22:69–75.

59. Pedersen J, Liang X, Casares-Magaz O, et al. Multivariate normal tissue complication probability models for rectal and bladder morbidity in prostate cancer patients treated with proton therapy. Radiother Oncol. 2020;153:279–288.

60. Luna JM, Chao HH, Diffenderfer ES, et al. Predicting radiation pneumonitis in locally advanced stage II–III non-small cell lung cancer using machine learning. Radiother Oncol. 2019;133:106–112.

61. Otten ML, Mikell CB, Youngerman BE, et al. Motor deficits correlate with resting state motor network connectivity in patients with brain tumours. Brain. 2012;135(4):1017–1026.

