## Supplemental tables and figures for "Fine Motor Skill Decline after Brain RT – A Multivariate Normal Tissue Complication Probability Study of a Prospective Trial"

| | Decision Stump | | Logistic Regression | | Spearman Correlation | | Non-parametric |
|---|---|---|---|---|---|---|---|
| | Cutoff | AUC (95% CI) | OR (95% CI) | p-value | $R_s$ | p-value | p-value |
| **Corticospinal Tract** | | | | | | | |
| $D_{max}$ | 19.61 | 0.60 (0.47 - 0.74) | 1.01 (0.97 - 1.04) | 0.64 | 0.09 | 0.57 | 0.29 |
| $D_{mean}$ | 0.57 | 0.52 (0.48 - 0.57) | 1.01 (0.97 - 1.06) | 0.60 | 0.07 | 0.65 | 0.33 |
| **Precentral Cortex** | | | | | | | |
| $D_{max}$ | 52.17 | 0.59 (0.47 - 0.71) | 1.00 (0.97 - 1.03) | 1.00 | 0.04 | 0.83 | 0.42 |
| $D_{mean}$ | 17.14 | 0.60 (0.46 - 0.74) | 0.98 (0.93 - 1.04) | 0.56 | $-0.07$ | 0.68 | 0.67 |
| **Precentral WM** | | | | | | | |
| $D_{max}$ | 27.63 | 0.58 (0.43 - 0.74) | 1.00 (0.97 - 1.03) | 0.96 | 0.01 | 0.93 | 0.47 |
| $D_{mean}$ | 0.31 | 0.58 (0.46 - 0.69) | 0.99 (0.94 - 1.04) | 0.64 | $-0.04$ | 0.82 | 0.60 |
| **Postcentral Cortex** | | | | | | | |
| $D_{max}$ | 57.54 | 0.57 (0.45 - 0.68) | 1.00 (0.98 - 1.03) | 0.78 | 0.03 | 0.85 | 0.43 |
| $D_{mean}$ | 0.12 | 0.58 (0.47 - 0.68) | 1.01 (0.95 - 1.06) | 0.85 | $<-.001$ | 0.98 | 0.52 |
| **Postcentral WM** | | | | | | | |
| $D_{max}$ | 58.95 | 0.57 (0.49 - 0.65) | 1.01 (0.98 - 1.03) | 0.67 | 0.10 | 0.53 | 0.27 |
| $D_{mean}$ | 0.10 | 0.58 (0.47 - 0.68) | 1.00 (0.96 - 1.06) | 0.88 | 0.03 | 0.86 | 0.43 |
| **Paracentral Cortex** | | | | | | | |
| $D_{max}$ | 37.17 | 0.61 (0.46 - 0.76) | 0.99 (0.96 - 1.01) | 0.37 | $-0.07$ | 0.68 | 0.67 |
| $D_{mean}$ | 22.70 | 0.60 (0.47 - 0.74) | 0.99 (0.95 - 1.02) | 0.52 | $-0.06$ | 0.71 | 0.65 |
| **Paracentral WM** | | | | | | | |
| $D_{max}$ | 33.16 | 0.61 (0.46 - 0.76) | 0.99 (0.96 - 1.02) | 0.48 | $-0.09$ | 0.59 | 0.71 |
| $D_{mean}$ | 0.24 | 0.58 (0.45 - 0.71) | 0.99 (0.95 - 1.03) | 0.60 | $-0.04$ | 0.83 | 0.59 |
| **Caudate** | | | | | | | |
| $D_{max}$ | 0.35 | 0.52 (0.48 - 0.57) | 1.00 (0.97 - 1.03) | 0.97 | 0.01 | 0.94 | 0.47 |
| $D_{mean}$ | 34.29 | 0.58 (0.46 - 0.69) | 0.99 (0.95 - 1.03) | 0.68 | $-0.05$ | 0.74 | 0.64 |
| **Pallidum** | | | | | | | |
| $D_{max}$ | 5.56 | 0.59 (0.47 - 0.71) | 1.00 (0.97 - 1.03) | 0.95 | $-0.03$ | 0.88 | 0.57 |
| $D_{mean}$ | 0.61 | 0.55 (0.48 - 0.61) | 1.01 (0.98 - 1.04) | 0.66 | 0.02 | 0.92 | 0.46 |
| **Putamen** | | | | | | | |
| $D_{max}$ | 1.53 | 0.55 (0.48 - 0.61) | 1.00 (0.97 - 1.02) | 0.77 | $-0.06$ | 0.70 | 0.66 |
| $D_{mean}$ | 0.32 | 0.55 (0.48 - 0.61) | 1.01 (0.97 - 1.04) | 0.76 | $-0.03$ | 0.84 | 0.59 |
| **Thalamus** | | | | | | | |
| $D_{max}$ | 0.85 | 0.55 (0.48 - 0.61) | 1.01 (0.98 - 1.04) | 0.61 | 0.07 | 0.65 | 0.33 |
| $D_{mean}$ | 21.37 | 0.59 (0.43 - 0.74) | 1.01 (0.98 - 1.04) | 0.63 | 0.08 | 0.61 | 0.31 |
| **Cerebellum Cortex** | | | | | | | |
| $D_{max}$ | 0.03 | 0.62 (0.53 - 0.72) | 1.00 (0.98 - 1.03) | 0.77 | 0.10 | 0.56 | 0.28 |
| $D_{mean}$ | 0.00 | 0.62 (0.53 - 0.72) | 0.99 (0.93 - 1.04) | 0.62 | 0.05 | 0.76 | 0.38 |
| **Cerebellum WM** | | | | | | | |
| $D_{max}$ | 0.01 | 0.60 (0.51 - 0.69) | 1.01 (0.98 - 1.04) | 0.68 | 0.10 | 0.54 | 0.27 |
| $D_{mean}$ | 0.00 | 0.60 (0.51 - 0.69) | 0.99 (0.94 - 1.04) | 0.70 | 0.07 | 0.66 | 0.34 |
| **Demographic/Clinical** | | | | | | | |
| Adjuvant Chemotherapy | – | 0.57 (0.42 - 0.72) | 1.82 (0.53 - 6.55) | 0.35 | 0.15 | 0.36 | 0.53 |
| Age | 27.00 | 0.57 (0.47 - 0.67) | 1.00 (0.96 - 1.05) | 0.95 | 0.04 | 0.80 | 0.40 |
| Anti-Epileptic Drug Use | – | 0.57 (0.42 - 0.72) | 1.79 (0.53 - 6.29) | 0.35 | 0.14 | 0.37 | 0.54 |
| Concurrent Chemotherapy | – | 0.55 (0.39 - 0.70) | 1.47 (0.44 - 5.06) | 0.54 | 0.10 | 0.55 | 0.76 |
| Diagnosis (Other) | – | 0.55 (0.40 - 0.70) | 0.67 (0.19 - 2.33) | 0.53 | $-0.10$ | 0.54 | 0.75 |
| Dose | 60.60 | 0.55 (0.48 - 0.61) | 1.09 (0.94 - 1.29) | 0.28 | 0.11 | 0.49 | 0.24 |
| Ethnicity (Non-Hispanic) | – | 0.57 (0.49 - 0.65) | – | 0.99 | 0.28 | 0.08 | 0.23 |
| Handedness (R) | – | 0.52 (0.42 - 0.62) | 0.63 (0.08 - 4.24) | 0.64 | $-0.07$ | 0.64 | 1.00 |
| KPS ($\leq$ 80) | – | 0.52 (0.44 - 0.60) | 2.11 (0.19 - 47.52) | 0.56 | 0.09 | 0.56 | 1.00 |
| Progression at 6mo | – | 0.57 (0.49 - 0.65) | – | 0.99 | $-0.28$ | 0.08 | 0.23 |
| Race (Other) | – | 0.50 (0.50 - 0.50) | 1.00 (0.11 - 9.05) | 1.00 | 0.00 | 1.00 | 1.00 |
| Radiation Modality (Protons) | – | 0.50 (0.50 - 0.50) | 1.00 (0.27 - 3.66) | 1.00 | 0.00 | 1.00 | 1.00 |
| Seizures | – | 0.62 (0.47 - 0.77) | 2.75 (0.79 - 10.38) | 0.12 | 0.24 | 0.12 | 0.21 |
| Sex (M) | – | 0.52 (0.37 - 0.68) | 1.22 (0.35 - 4.26) | 0.75 | 0.05 | 0.76 | 1.00 |
| Steroid Use | – | 0.52 (0.37 - 0.68) | 1.22 (0.35 - 4.26) | 0.75 | 0.05 | 0.76 | 1.00 |
| Surgery (any) | – | 0.55 (0.44 - 0.66) | 2.24 (0.39 - 17.63) | 0.39 | 0.14 | 0.39 | 0.66 |
| Surgery (GTR) | – | 0.52 (0.40 - 0.65) | 0.75 (0.16 - 3.34) | 0.71 | $-0.06$ | 0.72 | 1.00 |
| Tumor Laterality | – | 0.52 (0.37 - 0.68) | 1.21 (0.36 - 4.16) | 0.76 | 0.05 | 0.76 | 1.00 |

Table S1: Univariate analyses for decline on PNDH

| | Decision Stump | | Logistic Regression | | Spearman Correlation | | Non-parametric |
|---|---|---|---|---|---|---|---|
| | Cutoff | AUC (95% CI) | OR (95% CI) | p-value | $R_s$ | p-value | p-value |
| **Corticospinal Tract** | | | | | | | |
| $D_{max}$ | 59.33 | 0.59 (0.47 - 0.71) | 1.02 (0.98 - 1.07) | 0.31 | 0.17 | 0.27 | 0.14 |
| $D_{mean}$ | 7.28 | 0.63 (0.50 - 0.75) | 0.99 (0.93 - 1.04) | 0.65 | −0.04 | 0.81 | 0.60 |
| **Precentral Cortex** | | | | | | | |
| $D_{max}$ | 1.00 | 0.68 (0.52 - 0.84) | 1.00 (0.96 - 1.03) | 0.79 | −0.07 | 0.67 | 0.67 |
| $D_{mean}$ | 0.35 | 0.68 (0.52 - 0.84) | 1.01 (0.95 - 1.07) | 0.70 | −0.13 | 0.40 | 0.81 |
| **Precentral WM** | | | | | | | |
| $D_{max}$ | 0.78 | 0.68 (0.52 - 0.84) | 1.00 (0.97 - 1.03) | 1.00 | −0.05 | 0.77 | 0.62 |
| $D_{mean}$ | 42.45 | 0.59 (0.47 - 0.71) | 1.01 (0.96 - 1.07) | 0.57 | −0.09 | 0.55 | 0.73 |
| **Postcentral Cortex** | | | | | | | |
| $D_{max}$ | 0.05 | 0.59 (0.47 - 0.71) | 1.00 (0.96 - 1.03) | 0.82 | −0.11 | 0.48 | 0.77 |
| $D_{mean}$ | 0.00 | 0.59 (0.47 - 0.71) | 0.99 (0.93 - 1.05) | 0.84 | −0.15 | 0.34 | 0.84 |
| **Postcentral WM** | | | | | | | |
| $D_{max}$ | 9.79 | 0.59 (0.47 - 0.71) | 1.00 (0.97 - 1.04) | 0.87 | −0.04 | 0.78 | 0.62 |
| $D_{mean}$ | 2.29 | 0.59 (0.47 - 0.71) | 1.00 (0.94 - 1.05) | 0.97 | −0.12 | 0.46 | 0.78 |
| **Paracentral Cortex** | | | | | | | |
| $D_{max}$ | 0.45 | 0.71 (0.55 - 0.88) | 1.00 (0.97 - 1.03) | 1.00 | −0.09 | 0.58 | 0.72 |
| $D_{mean}$ | 0.30 | 0.71 (0.55 - 0.88) | 1.01 (0.97 - 1.05) | 0.73 | −0.11 | 0.48 | 0.77 |
| **Paracentral WM** | | | | | | | |
| $D_{max}$ | 0.51 | 0.71 (0.55 - 0.88) | 1.00 (0.97 - 1.03) | 0.88 | −0.07 | 0.64 | 0.69 |
| $D_{mean}$ | 0.33 | 0.69 (0.53 - 0.86) | 1.01 (0.97 - 1.06) | 0.55 | −0.08 | 0.59 | 0.71 |
| **Caudate** | | | | | | | |
| $D_{max}$ | 0.35 | 0.55 (0.46 - 0.63) | 1.00 (0.96 - 1.03) | 0.82 | −0.02 | 0.89 | 0.56 |
| $D_{mean}$ | 1.63 | 0.62 (0.46 - 0.78) | 0.99 (0.95 - 1.03) | 0.68 | −0.13 | 0.41 | 0.80 |
| **Pallidum** | | | | | | | |
| $D_{max}$ | 61.38 | 0.55 (0.46 - 0.63) | 1.00 (0.97 - 1.03) | 0.94 | −0.02 | 0.89 | 0.56 |
| $D_{mean}$ | 0.40 | 0.55 (0.46 - 0.63) | 0.99 (0.95 - 1.03) | 0.62 | −0.04 | 0.78 | 0.61 |
| **Putamen** | | | | | | | |
| $D_{max}$ | 62.08 | 0.55 (0.46 - 0.63) | 1.00 (0.97 - 1.04) | 0.85 | 0.03 | 0.85 | 0.43 |
| $D_{mean}$ | 6.65 | 0.66 (0.49 - 0.83) | 0.99 (0.95 - 1.03) | 0.72 | −0.07 | 0.66 | 0.68 |
| **Thalamus** | | | | | | | |
| $D_{max}$ | 3.14 | 0.62 (0.46 - 0.78) | 0.99 (0.96 - 1.02) | 0.65 | −0.06 | 0.70 | 0.66 |
| $D_{mean}$ | 1.26 | 0.63 (0.48 - 0.79) | 0.98 (0.94 - 1.02) | 0.44 | −0.13 | 0.40 | 0.81 |
| **Cerebellum Cortex** | | | | | | | |
| $D_{max}$ | 30.15 | 0.67 (0.55 - 0.80) | 0.98 (0.94 - 1.01) | 0.18 | −0.14 | 0.36 | 0.83 |
| $D_{mean}$ | 12.54 | 0.67 (0.55 - 0.80) | 0.96 (0.88 - 1.03) | 0.30 | −0.15 | 0.34 | 0.84 |
| **Cerebellum WM** | | | | | | | |
| $D_{max}$ | 21.38 | 0.66 (0.51 - 0.81) | 0.98 (0.94 - 1.01) | 0.22 | −0.14 | 0.37 | 0.82 |
| $D_{mean}$ | 12.19 | 0.64 (0.50 - 0.79) | 0.96 (0.89 - 1.02) | 0.26 | −0.14 | 0.38 | 0.82 |
| **Demographic/Clinical** | | | | | | | |
| Adjuvant Chemotherapy | – | 0.62 (0.45 - 0.79) | 0.37 (0.09 - 1.44) | 0.16 | −0.22 | 0.16 | 0.18 |
| Age | 41.50 | 0.71 (0.59 - 0.83) | 1.05 (1.00 - 1.11) | 0.05 | 0.29 | 0.06 | 0.03 |
| Anti-Epileptic Drug Use | – | 0.59 (0.42 - 0.76) | 0.49 (0.12 - 1.86) | 0.30 | −0.16 | 0.30 | 0.33 |
| Concurrent Chemotherapy | – | 0.66 (0.50 - 0.81) | 0.26 (0.05 - 1.05) | 0.07 | −0.28 | 0.07 | 0.09 |
| Diagnosis (Other) | – | 0.64 (0.47 - 0.80) | 3.08 (0.80 - 12.83) | 0.11 | 0.25 | 0.11 | 0.16 |
| Dose | 65.60 | 0.54 (0.46 - 0.62) | 0.94 (0.79 - 1.11) | 0.50 | −0.18 | 0.25 | 0.88 |
| Ethnicity (Non-Hispanic) | – | 0.51 (0.42 - 0.60) | 0.73 (0.06 - 16.70) | 0.81 | −0.04 | 0.81 | 1.00 |
| Handedness (R) | – | 0.52 (0.42 - 0.62) | 1.57 (0.20 - 32.57) | 0.70 | 0.06 | 0.71 | 1.00 |
| KPS ($\leq 80$) | – | 0.55 (0.50 - 0.60) | – | 0.99 | −0.17 | 0.28 | 0.55 |
| Progression at 6mo | – | 0.55 (0.50 - 0.60) | – | 0.99 | −0.17 | 0.28 | 0.55 |
| Race (Other) | – | 0.56 (0.50 - 0.62) | – | 0.99 | −0.19 | 0.21 | 0.56 |
| Radiation Modality (Protons) | – | 0.62 (0.46 - 0.79) | 3.00 (0.75 - 12.45) | 0.12 | 0.24 | 0.12 | 0.15 |
| Seizures | – | 0.55 (0.39 - 0.72) | 0.64 (0.15 - 2.49) | 0.53 | −0.09 | 0.54 | 0.73 |
| Sex (M) | – | 0.66 (0.50 - 0.82) | 0.26 (0.06 - 1.02) | 0.06 | −0.29 | 0.06 | 0.09 |
| Steroid Use | – | 0.65 (0.51 - 0.79) | 0.23 (0.03 - 1.04) | 0.08 | −0.28 | 0.07 | 0.09 |
| Surgery (any) | – | 0.56 (0.42 - 0.70) | 2.33 (0.40 - 12.67) | 0.32 | 0.15 | 0.32 | 0.37 |
| Surgery (GTR) | – | 0.66 (0.57 - 0.74) | – | 0.99 | 0.33 | 0.03 | 0.04 |
| Tumor Laterality | – | 0.53 (0.36 - 0.70) | 0.78 (0.20 - 2.99) | 0.71 | −0.06 | 0.72 | 0.75 |

Table S2: Univariate analyses for decline on DKEFS-TM

| |
|---|
| **Precentral cortex** |
| **Postcentral cortex** |
| **Paracentral cortex** |
| **Precentral superficial white matter** |
| **Postcentral superficial white matter** |
| **Paracentral superficial white matter** |
| **Corticospinal tract** |
| **Cerebellar cortex** |
| **Cerebellar superficial white matter** |
| **Thalamus** |
| **Caudate** |
| **Putamen** |
| **Pallidum** |

Table S3: Motor-associated regions of interest

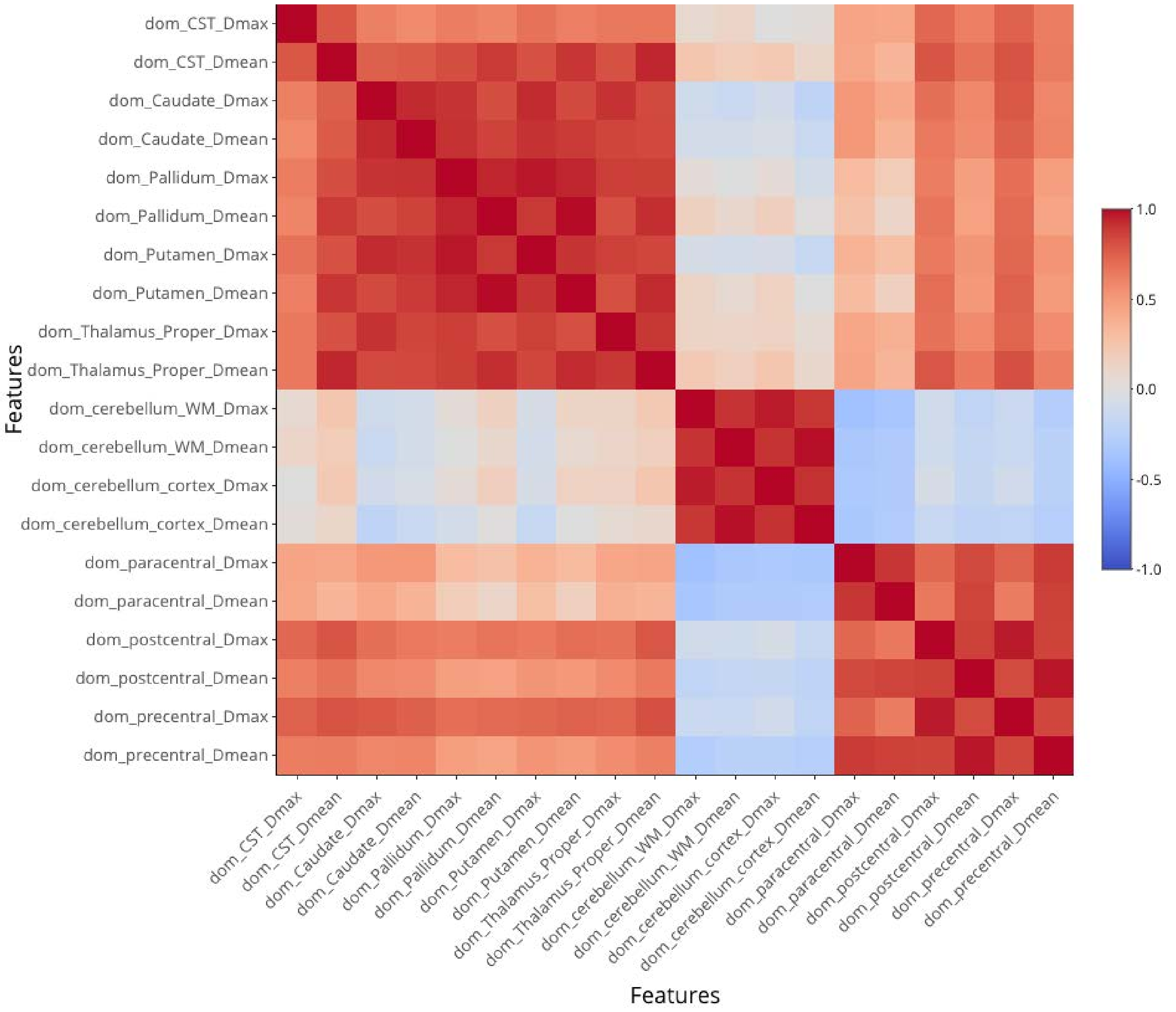


Fig S1: Pearson correlation heat map for dominant-sided dosimetric variables.

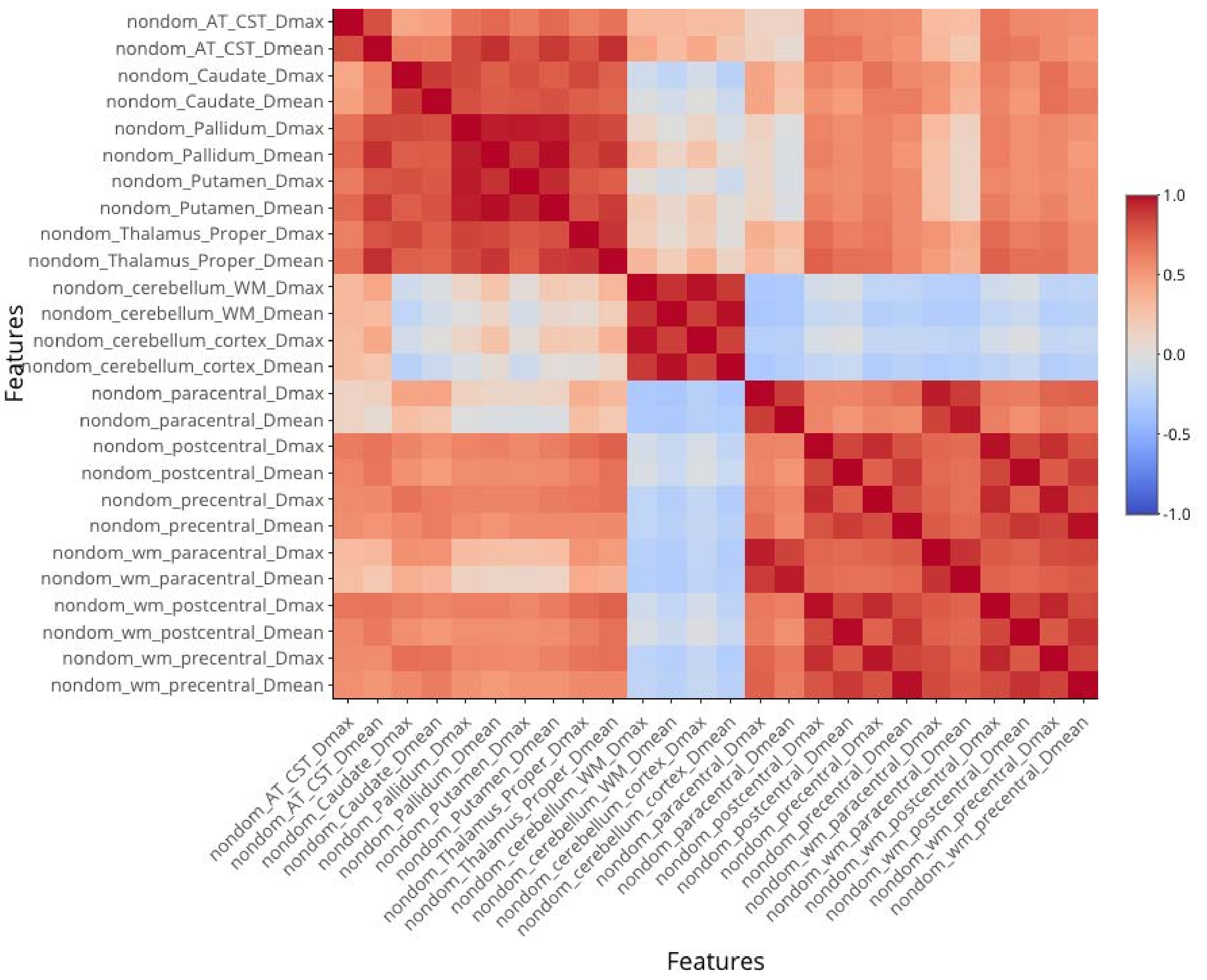


Fig S2: Heatmap for Pearson correlation between non-dominant sided dosimetric variables

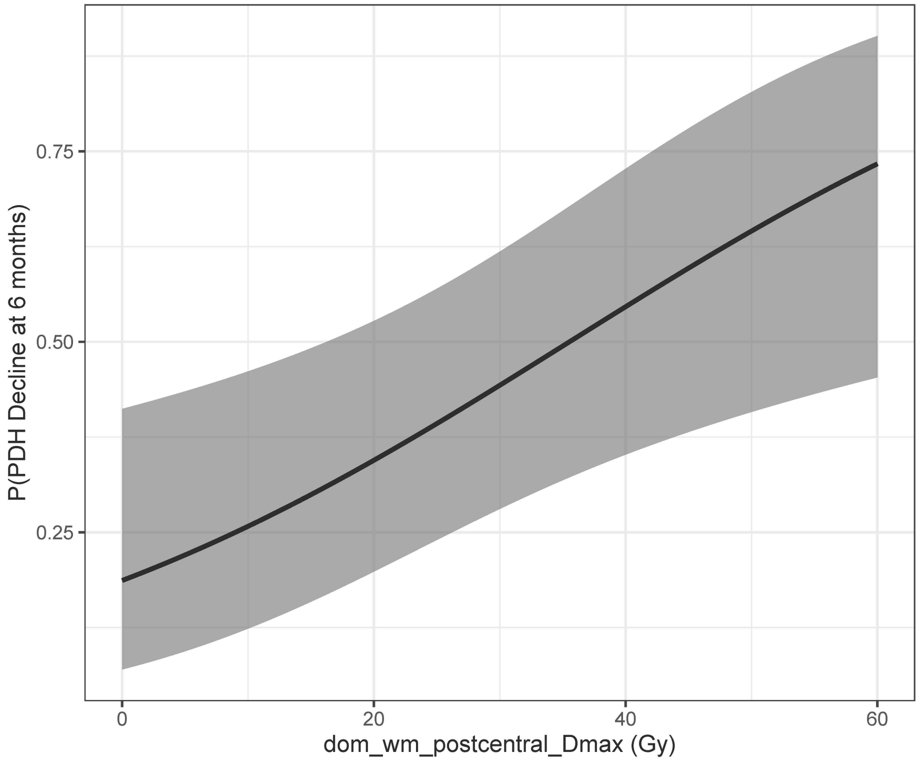


Fig S3: Logistic NTCP curve for the single term model produced using automated bootstrapped regression

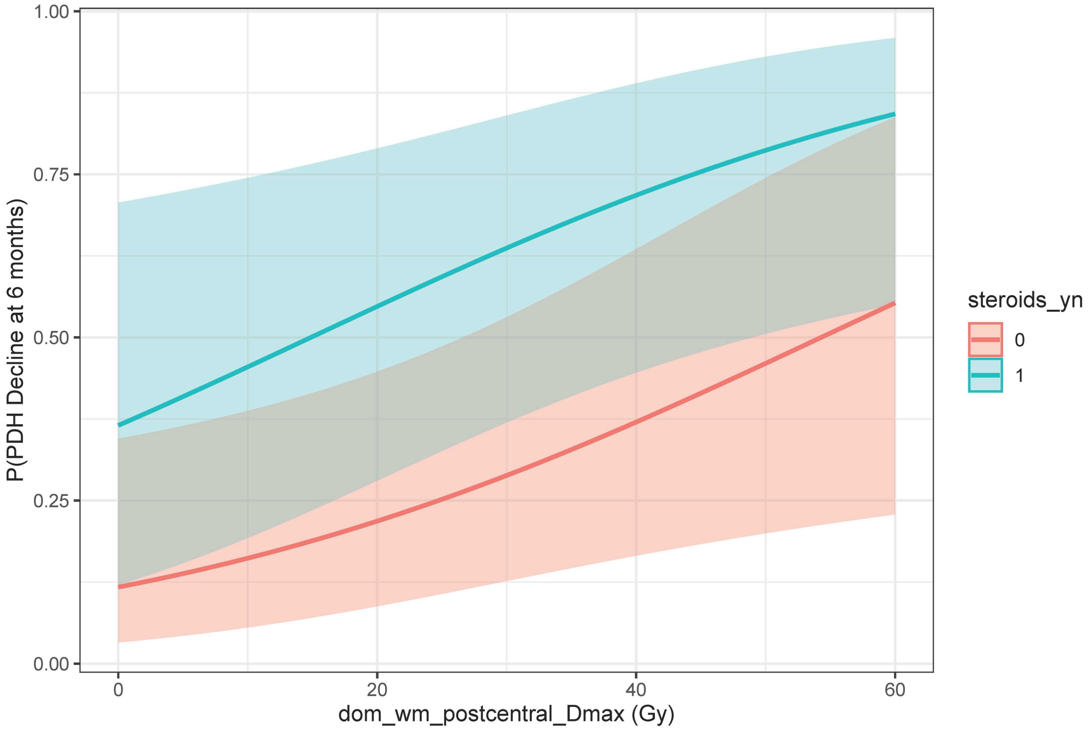


Fig S4: Logistic NTCP curve for the two term model produced using LASSO (coefficients here are unregularized)
